# A SEIRD+V Model for the Effect of Vaccination and Social Distancing on SARS-CoV-2 Infection and Mortality

**DOI:** 10.1101/2022.10.23.22281416

**Authors:** Alexander Jin, Husham Sharifi

## Abstract

We present a deterministic, calibrated Susceptible-Exposed-Infected-Recovered-Dead + Vaccinated (SEIRD+V) model that simulates the spread and containment of COVID-19. We use the model to compare the effectiveness of vaccination vs. social distancing on death prevention and total and peak infection reduction. We find that vaccination drastically reduces total deaths from COVID-19, as well as total and peak infections with Severe Acute Respiratory Syndrome Coronavirus 2 (SARS-CoV-2). We find that social distancing plays a role in reducing total COVID-19 deaths, but its impact is less pronounced when vaccine efficacy and vaccination fraction are both high. Social distancing also plays a role in reducing total and peak infections, which is attenuated in the presence of vaccination. We employed a thresholding methodology to assess the requirements of vaccine efficacy and the vaccination fraction to limit total COVID-19 deaths and peak infections to a 5% threshold. Our thresholding results quantify the impact of social distancing on total COVID-19 deaths and peak infections and are significant in their ability to inform public health policy for future outbreaks, as well as for SARS-CoV-2 itself as it continues to mutate and alter its transmissibility.

## 1 Introduction

SARS-CoV-2 was first discovered and identified in Wuhan, China, in December 2019 and shortly thereafter spread around the globe. As of this writing, there are have been 580 million documented cases of infection and 6.41 million recorded deaths (1). We continue to experience in certain countries quarantine and isolation protocols, mandatory work furloughs for infected individuals, and modified arrangements for distancing in educational environments such as secondary schools. National governments also have devised containment policies, and the Centers for Disease Control (CDC) in the United States (US) has explicit policies for quarantine and isolation (2). These measures and their efficacy hold relevance for our ongoing efforts to combat transmission during times of lower prevalence, during any significant recrudescence of the virus, and during future respiratory pandemics from alternative viruses. In parallel scientific communities around the globe have developed vaccines to combat COVID-19 at unprecedented speed and with broad, rapid distribution. Together, these policies and medical interventions provide a composite public health approach to mitigating the health impact of Severe Acute Respiratory Syndrome Coronavirus 2 (SARS-CoV-2) on individuals and communities.

As health experts and government authorities formulate regulatory policies, mathematical models have proven helpful for investigating the spread of the virus through theoretical frameworks that showcase the effectiveness of various intervention strategies and the impact of vaccination on protecting populations from viruses (3,4). They can provide public health officials and the general public with insights and predictions pertaining to the spread of pathogens and the effectiveness of containment measures and vaccination that recorded data alone cannot (5,6). The role of mathematical modelling in improving public health responses to infectious pandemics has been supported by the CDC Infectious Disease Modelling and Analytics Initiative (7).

This is an especially significant concern in that climate change and loss of biodiversity is anticipated to increase the frequency of new pandemics (8,9), with pandemic risk projected to increase by three-fold in the next few decades (10). Given heightened risk and increased frequency of new pandemics, preparedness through modelling can be a key component for public health response systems. Mathematical modelling can help preparedness by advancing technical knowledge and techniques during existing pandemics that can be applied forward as new pandemics emerge.

Of pandemics mathematical models, the Susceptible-Exposed-Infected-Recovered (SEIR) model has been widely used to model disease transmission dynamics in epidemics (e.g., tuberculosis and varicella) (11,12), and other mathematical models have been used in a number of studies for COVID-19 (13–16). In one study Carcione and colleagues applied SEIR to simulate the COVID-19 epidemic in the Lombardy Region of Italy (17). By use of the same data in the Carcione study, we developed an enhanced SEIR model with modifications to improve performance by inclusion of the following: 1. effect of vaccinations on total deaths and peak infections from COVID-19; 2. Vaccine efficacy; 3. Interactions between vaccine efficacy and vaccine rate; and 4. A thresholding methodology to assess minimum requirements that are needed to satisfy pre-set goals (e.g., 5% rate of peak infection). These enhancements have been demonstrated to be important in prior models but have not be applied in an integrated analytic model, nor have they been used for a model to assess a combination of vaccination and social distancing (18,19).

The paper is organized as follows. First, the SEIRD+V (Susceptible-Exposed-Infected-Recovered-Dead+Vaccinated) model is introduced, and its key aspects are discussed. Then the model is applied to the Lombardy dataset, with validity assessed by comparison to the Carcione study. Finally, the effects of social distancing and vaccination are, respectively, varied and incorporated into the model to assess the impact of social distancing and vaccination on the containment of COVID-19. Thresholding is applied to assess the requirements of vaccine efficacy and vaccination fraction to reach preset COVID-19 death and peak infection goals.

## 2 Methods and Materials

### 2.1 The Basic SEIRD Model

We first build a SEIRD (Susceptible, Exposed, Infected, Recovered, Dead) model, officially adding a heretofore implicit Dead compartment. See Figure 1. The model divides the total population N_0_ into five compartments at any given time: Susceptible (S), Exposed (E), Infectious (I), Recovered (R), and Dead (D).

**Figure 1.**
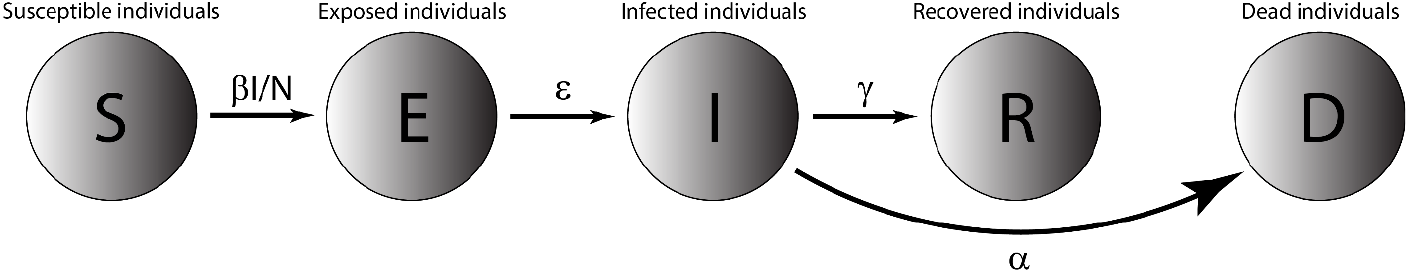
The basic SEIRD Model. The whole population is divided into 5 compartments: Susceptible, Exposed, Infected, Recovered, and Dead. β, *ϵ*, γ, and α are parameters characterizing transitions between the relevant compartments. These transition parameters are described below. Although there are technically links between the S, E, and R compartments to D (through natural deaths), adding all of them would clutter the diagram, so we have left them out.

- Susceptible (S): Individuals of the population who are not infected but can become infected. A susceptible individual becomes exposed when they come into contact with an infectious individual.
- Exposed (E): Individuals of the population who have come into contact with an infectious individual. Exposed individuals may become infected and carry the virus, but do not shed virus in sufficient density to infect other individuals. Not all exposed individuals become infected. Exposed individuals who are infected do not show symptoms, and the disease is latent in these individuals at this stage.
- Infectious (I): Individuals who have been infected by the virus and can transmit it to susceptible individuals. An infectious individual remains infectious for a period of time and is removed from the infectious population when they recover from disease or die. Note that naming of this compartment is not consistent in the literature. It is named “Infected” in some papers (e.g., 16,17) and “Infectious” in others (13,20). When referring to individuals in this compartment, this paper uses “infected” and “infectious” interchangeably.
- Recovered (R): Individuals who have recovered from the disease and are assumed to be immune.
- Dead (D): Individuals who have died (i.e., all-cause mortality).

The model assumes that the time scale of the model is short enough such that births and natural deaths (deaths not caused by COVID-19) are negligible and that the number of deaths from COVID-19 is small compared to the living population.

The ordinary differential equations that govern the transitions of the population from one compartment to the next are as follows:

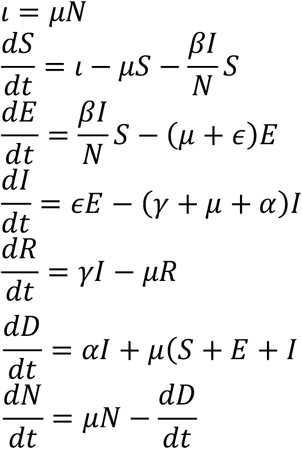

where *N = S + E + I + R ≤ N*_*0*_. Equations are subject to the initial conditions *S(0), E(0), I(0), R(0), and N(0)*.

The parameters are defined as:

𝜄: Birth rate.

μ: Per-capita natural death rate.

α: Virus-induced average fatality rate.

β: Probability of disease transmission per contact (dimensionless) times the number of contacts per unit time.

*ϵ*: Rate of progression from exposed to infectious (the reciprocal is the latent period).

γ: Recovery rate of infectious individuals (the reciprocal is the infectious period).

In the model, the choice 𝜄 = μ = 0 and *ϵ* = ∞ gives the classical SIR model, while if ε and μ are not zero, the model is termed an endemic SIR model (3). However, the SIR model has no latent stage (no exposed individuals), so it is, therefore, inappropriate as a model for diseases with an *ϵ* such as that of COVID-19.

### 2.2 The SEIRD model + Vaccination

This model extends our basic SEIRD model described in 2.1, by further dividing each compartment in the basic model based on vaccination status (except the Dead compartment). This model divides the total population N_0_ into nine compartments at any given time: Susceptible unvaccinated (S), Exposed unvaccinated (E), Infectious unvaccinated (I), Recovered unvaccinated (R), Susceptible vaccinated (SV), Exposed vaccinated (EV), Infectious vaccinated (IV), Recovered vaccinated (RV), and Dead (D).

- Susceptible unvaccinated (S): Unvaccinated individuals of the population who are not infected but can become infected, when coming into contact with an infectious individual (unvaccinated or vaccinated).
- Exposed unvaccinated (E): Unvaccinated individuals of the population who have come into contact with an infectious individual (unvaccinated or vaccinated).
- Infectious unvaccinated (I): Unvaccinated individuals who have been infected by the virus. Those individuals transmit the virus to unvaccinated and vaccinated susceptible individuals.
- Recovered unvaccinated (R): Unvaccinated individuals who have recovered from the disease.
- Recovered individuals are susceptible to be reinfected, albeit at lower probabilities.
- Susceptible vaccinated (S_V_): Vaccinated individuals of the population who are not infected but can become infected. Even though susceptible, the probabilities of vaccinated individuals being infected is much lower than unvaccinated individuals (21,22).
- Exposed vaccinated (E_V_): Vaccinated individuals of the population who have come into contact with an infectious individual (unvaccinated or vaccinated). Exposed vaccinated individuals are less likely to develop symptoms and are less infectious than exposed unvaccinated individuals.
- Infectious vaccinated (I_V_): Vaccinated individuals who have been infected. Vaccinated individuals are less infectious, less likely to develop severe symptoms or die than unvaccinated infectious individuals.
- Recovered vaccinated (R_V_): Vaccinated individuals who have been infected by the virus and recovered from the disease.
- Dead (D): Individuals who have died (from both the disease and natural causes)

This model is illustrated in Figure 2. It similarly assumes that the time scale of the model is short enough so that births and natural deaths (deaths not caused by COVID-19) are negligible and that the number of deaths from COVID-19 is small compared to the living population.

**Figure 2.**
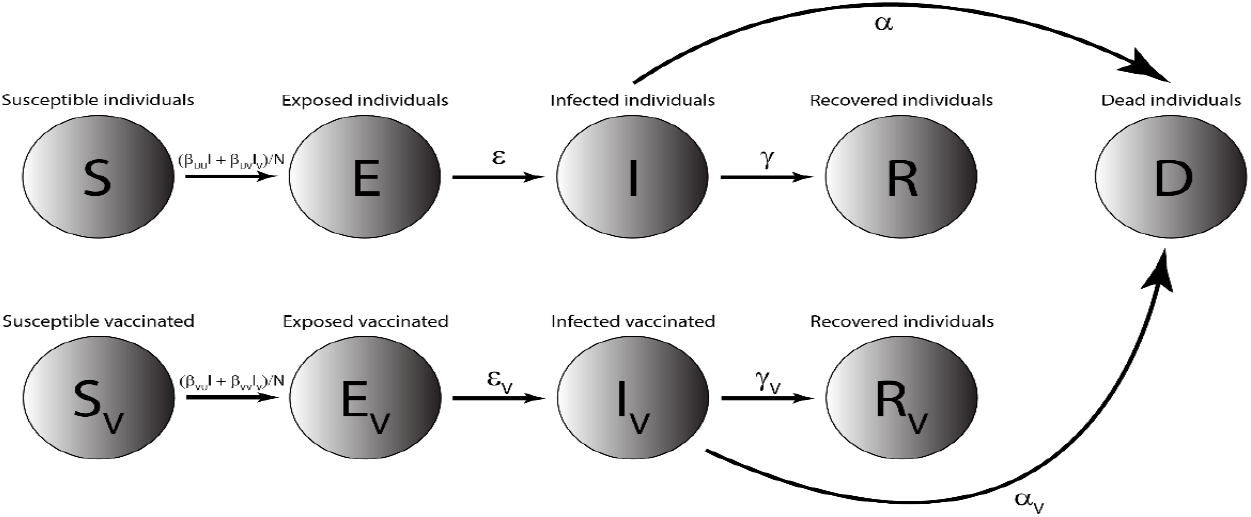
The SEIRD+V Compartmental Model. This model is enhanced from the basic SEIRD model in Figure 1, with each of the S, E, I, and R compartment further divided into an unvaccinated and a vaccinated compartment. The transition parameters are labelled on the arrows between the compartments. Although there are once again technically links between all living compartments to D (through natural deaths), adding all of them would clutter the diagram, so we have left them out.

The ordinary differential equations that govern the transitions of the population from one compartment to the next are as follows:

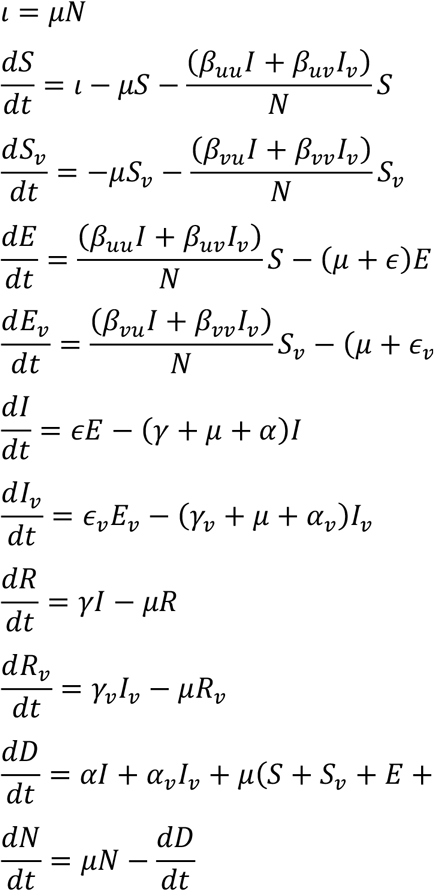

where *N = S + E + I + R + S*_*V*_ *+ E*_*V*_ *+ I*_*V*_ *+ R*_*V*_ *≤ N*_*0*_. Equations are subject to the initial conditions *S(0), E(0), I(0), R(0), S*_*V*_*(0), E*_*V*_*(0), I*_*V*_*(0), R*_*V*_*(0), and N(0)*. The parameters for the extended model are defined as:

𝜄: Birth rate.

*μ*: Per-capita natural death rate.

*α*: Virus-induced average fatality rate among unvaccinated individuals.

*α*_*v*_: Virus-induced average fatality rate among vaccinated individuals.

*β*_*uu*_: Probability of disease transmission per unvaccinated contact (dimensionless) times the number of unvaccinated contacts per unit time.

*β*_*ij*_: Probability of disease transmission per i contact (dimensionless) times the number of j contacts per unit time.

*β*_*vu*_: Probability of disease transmission per vaccinated contact (dimensionless) times the number of unvaccinated contacts per unit time.

*β*_*vv*_: Probability of disease transmission per vaccinated contact (dimensionless) times the number of vaccinated contacts per unit time.

*ϵ:* Rate of progression from exposed, unvaccinated to infectious, unvaccinated (the reciprocal is the incubation period).

*ϵ*_*vv*_: Rate of progression from exposed, vaccinated to infectious, vaccinated (the reciprocal is the incubation period).

*γ*: Recovery rate of infectious, unvaccinated individuals (the reciprocal is the infectious period).

*γ*_*v*_: Recovery rate of infectious, vaccinated individuals (the reciprocal is the infectious period).

We use the following parameters and initial conditions:

*κ*: Fraction of vaccinated population

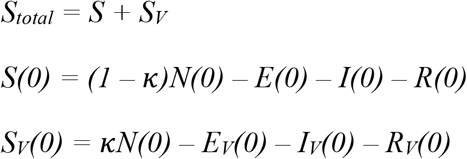

*ξ:* Vaccine efficacy

*ξ*_*uv*_ Vaccine efficacy in preventing an unvaccinated individual from infecting a vaccinated individual

*ξ*_*vu*_: Vaccine efficacy in preventing a vaccinated individual from infecting an unvaccinated individual

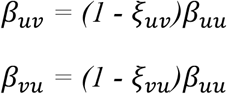

We assume that any possible *ξ*_*uv*_ ≈ *ξ*_*vu*_, so that *ξ*_*uv*_ *= ξ*_*vu*_ = *ξ*. Similarly, *β*_*vv*_: is set to *β*_*uv*_ × *β*_*vu*_.

### 2.3 Thresholding

The thresholding process takes a threshold input and calculates all parameter combinations where the specified threshold condition is met. We illustrate the threshold with a graph in which all coordinates corresponding to the parameter pairs where the threshold is crossed are identified by color, leaving blank all coordinates that do not cross the threshold.

For example, COVID death toll is dependent on vaccination rate *κ* and vaccine efficacy *ξ*. Model thresholding would take the death toll > 5% of some arbitrary value as a threshold input and then collect all the (*κ, ξ*) combinations under which the COVID death toll is greater than that 5%, coloring in their coordinates as the model computes. Spaces in the chart that are clear represent spaces where the COVID death toll is less than that 5%.

### 2.4 Datasets

Data for our model come from those used in the study by Carcione and colleagues, who processed and labeled model constraints and initial-final values from the initial months of epidemic data in Lombardy, Italy (23). The dataset covers 02/24/2020 to 05/05/2020 and includes the number of patients for the following variables on a per-day basis: hospitalized patients; intensive care patients; quarantined individuals; total individuals with positive tests; new positive tests; patients who were discharged from admission or recovered from infection; number of deaths; and number of tests administered (24). This dataset will be referred to as the Lombardy dataset.

## 3 Results

### 3.1 Validation of our models

We first tested our basic SEIRD model and our enhanced SEIRD+V model against the Lombardy dataset, using degenerate cases that simplify to Carcione’s model. We also replicated Carcione’s model with original code. We then applied all three models to the Lombardy dataset, with results shown in Figure 3. We used the same parameters from Carcione for all three models, only adding the vaccinated compartments to the first two models and splitting the susceptible population from Carcione between the unvaccinated and vaccinated susceptible compartments.

**Figure 3.**
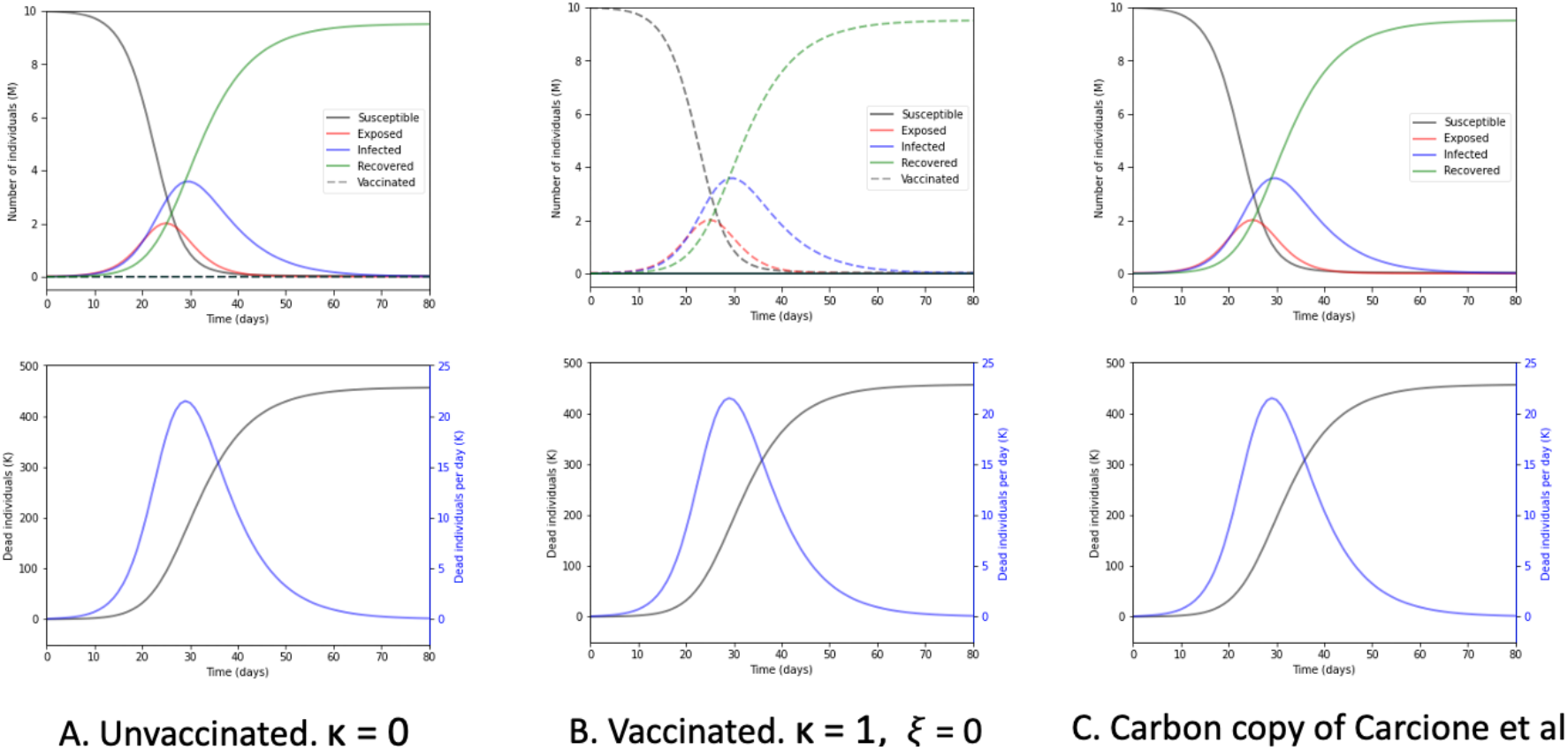
Comparison of simulation results. The three different models are applied to the same dataset (the Lombardy data set). A) Resultant graphs produced by our SEIRD+V model when *κ* = 0 and *ξ* = 0. B) Resultant graphs produced by the same model when *κ* = 1 and *ξ* = 0. C) Resultant graphs replicating Carcione et al’s model.

A comparison of these three sets of charts reveals that they are identical. Peak infections, peak deaths per day, maximum total deaths, and the shapes of their respective curves are all the same. The other curves (i.e., susceptible, exposed, recovered) are similarly identical between the charts.

### 3.2 Effects of vaccination on SEIRD+V dynamics

We next simulate the effect of vaccination on COVID infections, COVID deaths, and the dynamics of other compartments of the model, as shown in Figure 4.

**Figure 4.**
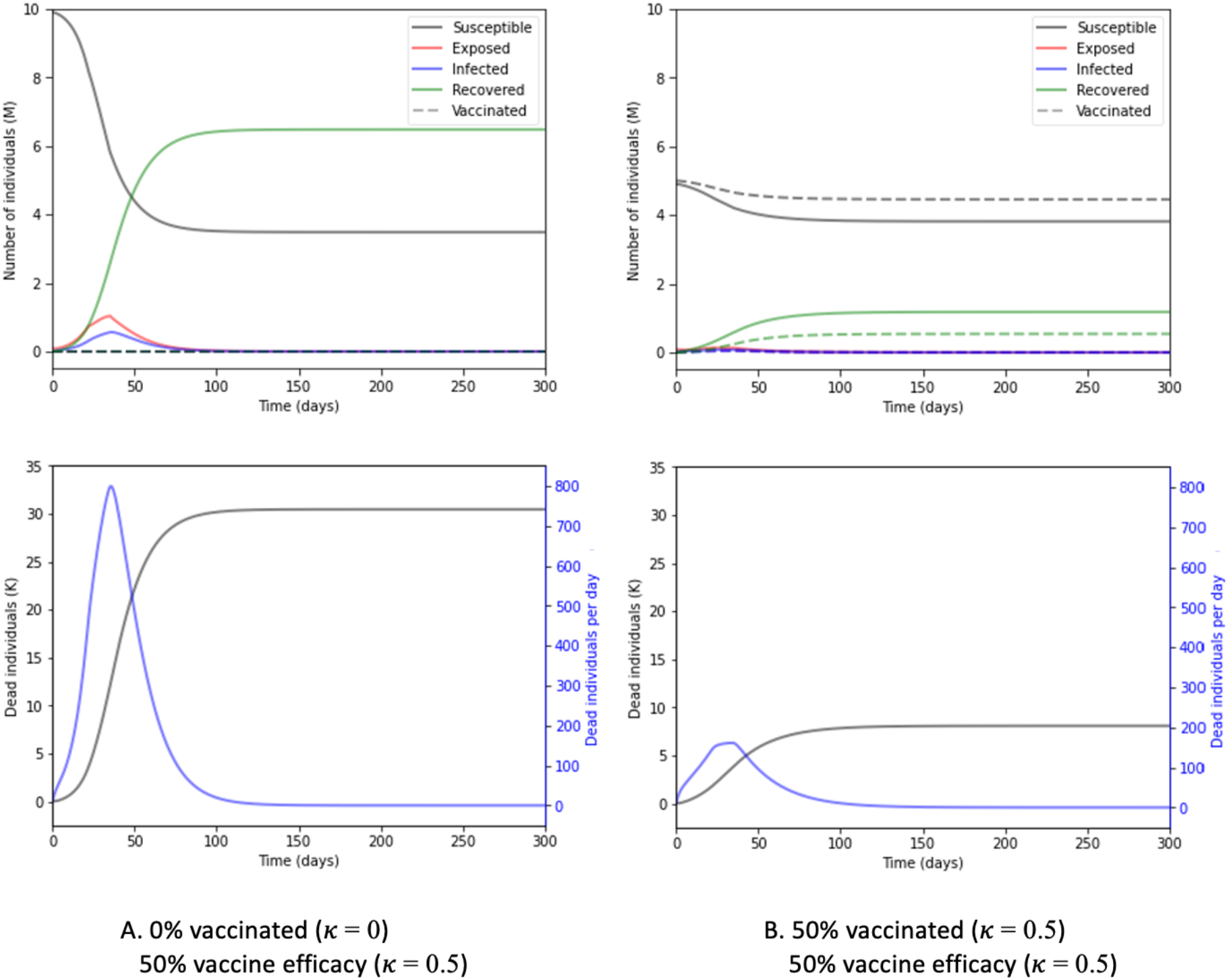
Effect of vaccination on SEIRD+V dynamics. A) 0% vaccinated *κ* = 0, 50% vaccine efficacy *ξ* = 0.5. 𝐵) 50% vaccinated *κ* = 0.5, 50% vaccine efficacy *ξ* = 0.5. Vaccine efficacies are the same for A and B, but A has 0% of the population vaccinated (*κ* = 0), while B has 50% of the population vaccinated (*κ* = 0.5). For brevity, the dashed line for vaccinated indicates that the dashed pattern applies to all curves that are generated in the vaccinated case.

In the examples above, we apply the model to a population with the following initial conditions and model parameters:

**Table 1.**
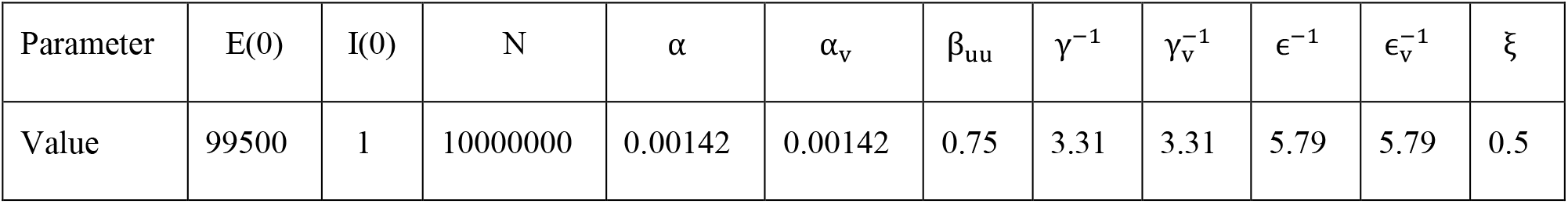
Initial conditions and model parameters.

| Parameter | $E(0)$ | $I(0)$ | $N$ | $\alpha$ | $\alpha_v$ | $\beta_{uu}$ | $\gamma^{-1}$ | $\gamma_v^{-1}$ | $\epsilon^{-1}$ | $\epsilon_v^{-1}$ | $\xi$ |
| --- | --- | --- | --- | --- | --- | --- | --- | --- | --- | --- | --- |
| Value | 99500 | 1 | 10000000 | 0.00142 | 0.00142 | 0.75 | 3.31 | 3.31 | 5.79 | 5.79 | 0.5 |

Comparison of Figure 4(A) and 4(B) reveals that vaccination significantly impact the dynamics of COVID-19. With 50% vaccine uptake rate of 50% efficacy vaccines, the number of susceptible people at the beginning of an epidemic is reduced by ∼55%. Interestingly, in later stages, the number of susceptible people are about even with or without vaccination. Note that in Figure 4(B), in later stages, the total number of susceptible individuals are twice that in Figure 4(A). This is because vaccinated people are mostly immune (i.e., not infected), and thus they remain susceptible. Most significant is the impact of vaccination on peak exposed and peak infected. Without vaccination, the number of peak exposed and that of peak infected around 1.038 million and 0.563 million, respectively. Vaccination drastically flattens both peaks. With vaccination, peak exposed and is 0.202 million (−80.5%) and peak infected 0.113 million (−79.8%). Vaccination also has a large impact on peak death rate and on total deaths. With vaccination, peak death rate and total deaths are reduced by 79.8% (from 799 to 161 deaths per day) and 73.5% (from 30,471 to 8080 deaths), respectively.

### 3.3 Effects of social distancing

Drawing from the Carcione model, we add a social distancing component by adding the parameter *β*_*ext*_, which is a list of time-dependent values for *β*_*uu*_ that work as follows:

When time t <= 22 days:

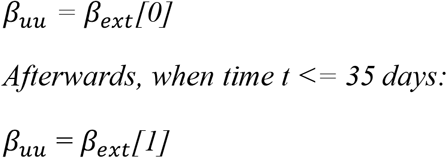

After time t exceeds 35 days,

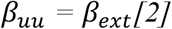

### 3.4 Model applications to Lombardy dataset: Total deceased

With social distancing in place, we next vary social distancing (yes/no), vaccination fraction *κ*, and vaccine efficacy *ξ* to see their effects on the total number of deceased individuals. The value of “Total deceased” is the final value of *D(t)* stored by the program after running the SEIRD+V simulation. Applying this to the Lombardy dataset produces results on the impact of vaccine efficacy and vaccination rate on total deceased with or without social distancing.

Figure 5(A) shows the total COVID deaths for different vaccination fractions of the population, with various values of vaccinated transmission probabilities, when there is no social distancing. For all vaccinated transmission probability values, the COVID death toll decreases as the vaccination fraction increases. There are no exceptions.

**Figure 5.**
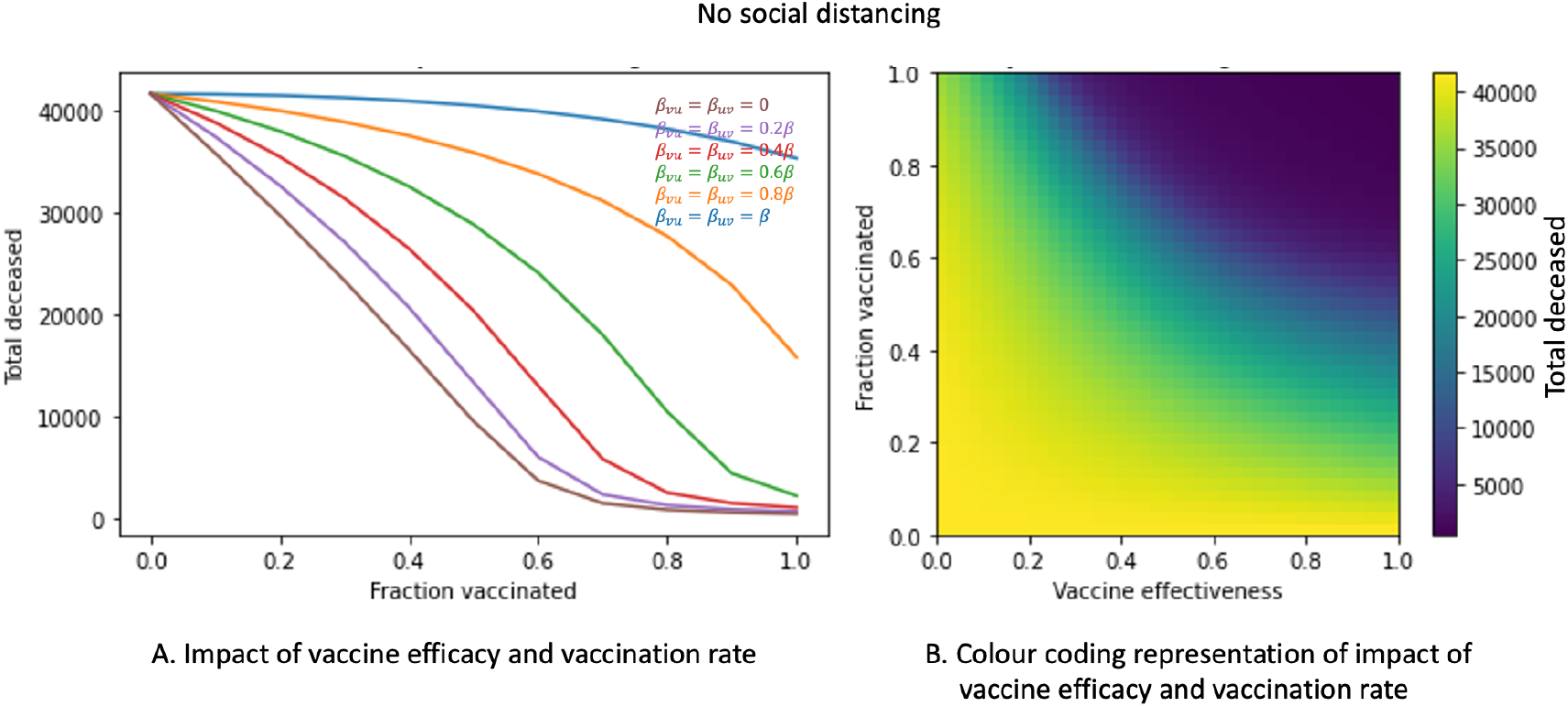
Impact of vaccine efficacy and the vaccination fraction on total COVID deaths, with no social distancing. A) Impact of vaccine efficacy and vaccination rate. B) Color coding representation of Impact of vaccine efficacy and vaccination rate. The unvaccinated to unvaccinated transmission probability *β*_*uu*_ = 0.75 for all time.

We model vaccine efficacy by vaccinated transmission probability. The left chart of Figure 5 also shows that total COVID deaths decline more sharply with the vaccination fractions as vaccinated transmission probability decreases. This result suggests that with increasing vaccine efficacy, the COVID death toll decreases for all vaccination fractions. This result is also represented in a color coding scheme in Figure 5(B). The color gradient on the right depicts the number of total COVID deaths, with darker blue shades representing fewer deaths and darker yellow shades representing more deaths, for varying combinations of *κ* and *ξ*. As vaccine efficacy moves from left to right, and the vaccination fraction from bottom to top, the shade of the color moves from yellow to green to dark blue, i.e., decreasing COVID deaths.

Figure 6 shows the same simulation as Figure 5, with social distancing added. It is clear that, with social distancing, the impact of vaccinated transmission probability on total COVID deaths is less pronounced than when there is no social distancing. Comparing Figure 6(A) to Figure 5(A), all the curves in Figure 6(A), left panel shift lower (i.e., decreased COVID death toll). However, the differences between the curves for the various *β*_*uv*_/*β*_*vu*_ values have narrowed. Figure 6, right panel, shows that the color spectrum has drastically shifted to the lower left corner, when compared with Figure 5, left panel.

**Figure 6.**
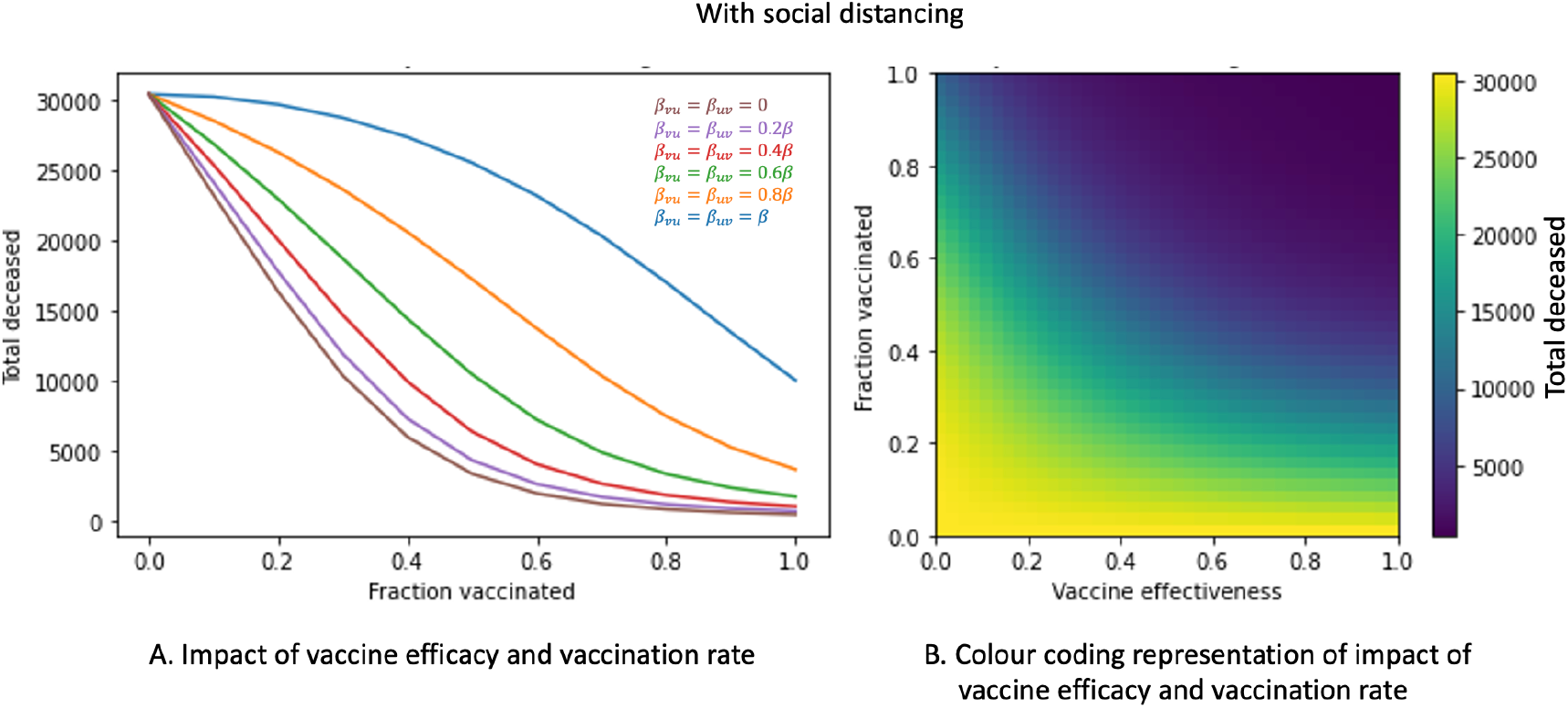
Impact of vaccine efficacy and the vaccination fraction on total COVID deaths, with social distancing. A) Impact of vaccine efficacy and vaccination rate. B) Color coding representation of Impact of vaccine efficacy and vaccination rate. *β*_*uu*_ changes at specific times to reflect social distancing adjustments: *β*_*uu*_ = 0.75 at t = 0, changes to *β*_*uu*_ = 0.57 at t = 22 days, and then changes to *β*_*uu*_ = 0.395 at t = 35 days.

As the above figures demonstrate, the death toll at *κ* = 0 is, for all *ξ* values (even *ξ* = 0), significantly larger than the death toll at *κ* = 1. The color-bars on the right depict the changing death toll in varying intensities of color as both *κ* and *ξ* are varied.

Figure 7 is the overlaid product of two thresholding simulations for total COVID deaths, one with social distancing (blue) and the other without social distancing (light blue). The threshold is set to less than 5% of the greatest observed COVID death toll in the simulation, 41,000. The first shades all (*κ, ξ*) combinations for which the death toll meets that thresholding condition (total COVID death > 5%) in the ‘distancing’ simulation, and the second shades the same combinations meeting the same thresholding condition in the ‘no distancing’ simulation. Simulation results indicate that with social distancing, vaccine efficacy must reach at least 39% and the vaccination fraction must reach at least 54% to attain the goal of <5% COVID death. Without social distancing, 5% COVID death threshold requires a little higher vaccine efficacy (56%) and vaccination fraction (97%). From the chart, it can be seen that the light blue (no social distancing) extends a little more to the upper right compared to the dark blue (social distancing).

**Figure 7.**
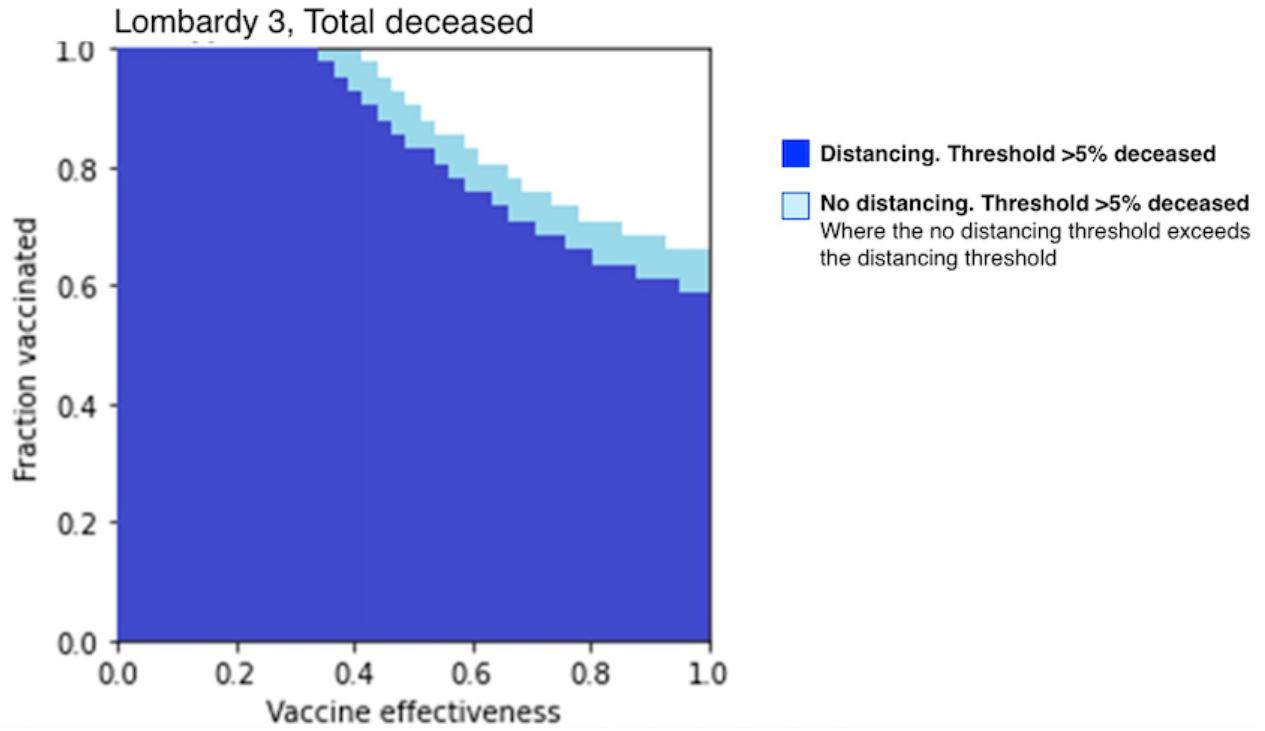
Thresholding of total COVID deaths, social distancing vs. without social distancing. In the graph, the clear area represents the vaccine efficacy/vaccination fraction combinations where total COVID deaths < 5%. Note that the clear area for blue is larger than that for light blue.

This type of overlaid thresholding according to prespecified percentage has applications pertaining to targeted reduction, as target percentages can be inputted into the simulation and used to compare vaccination thresholds for ‘distancing’ and ‘non-distancing’ populations. In this case, social distancing populations require less vaccinations and lesser vaccine efficacies to reach the same threshold of 5% of the target (41,000) compared to non-distancing populations.

### 3.5 Model applications to Lombardy data set: Peak infected

Next, we vary social distancing, *κ*, and *ξ* to see their effects on peak infections. The value of “Peak infected” is essentially just the maximum value of *I(t)* stored by the program after running the SEIRD+V simulation.

Analogous to Figures. 5 and 6, Figures 8 and 9 present the simulation results for the impact of vaccinated transmission probability (*β*_*vu*_) and the vaccination fraction (*κ*) but do so in this case on peak infections. Figure 8 presents the results when there is no social distancing; whereas, Figure 9 presents the results when there is social distancing.

**Figure 8.**
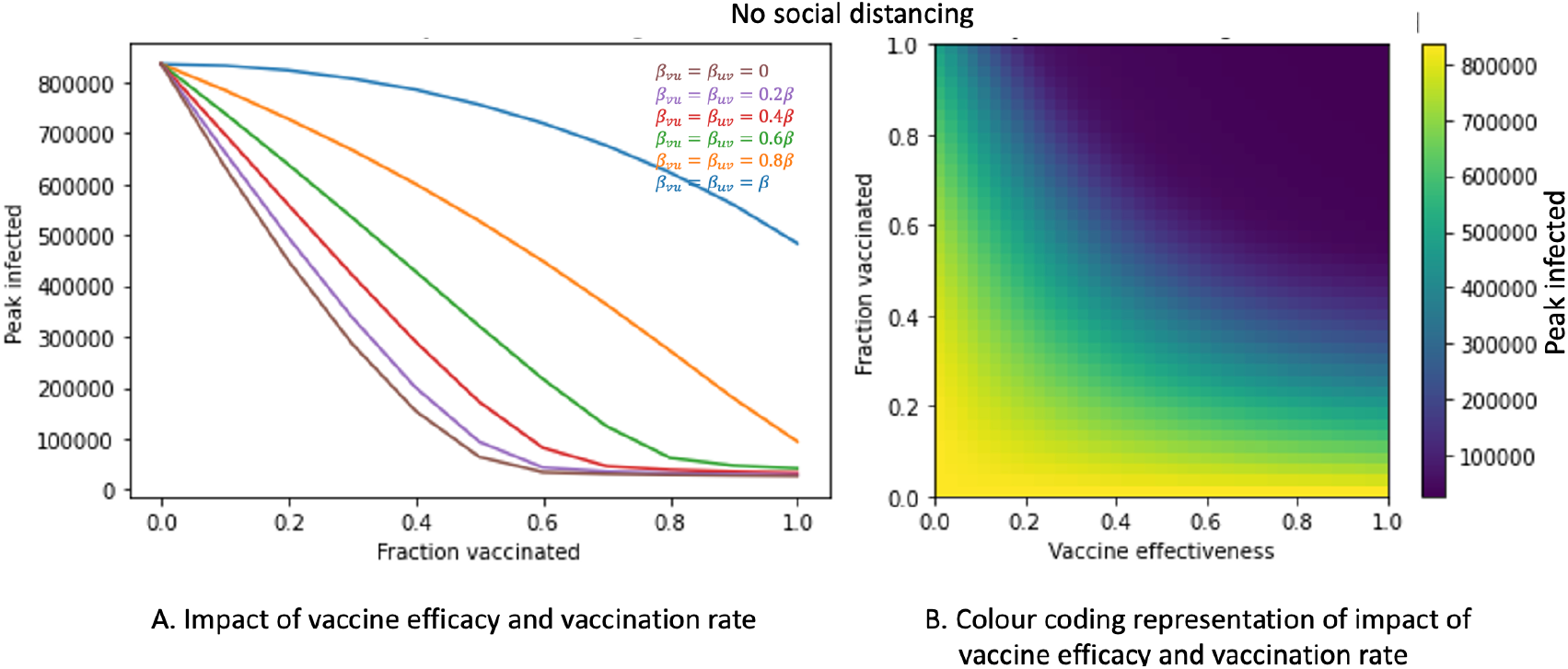
Impact of vaccine efficacy and vaccination fraction on peak COVID infections, with no social distancing. A) Impact of vaccine efficacy and vaccination rate. B) Color coding representation of Impact of vaccine efficacy and vaccination rate. This is similar to Figure 5, but with peak COVID infections as the study target.

**Figure 9.**
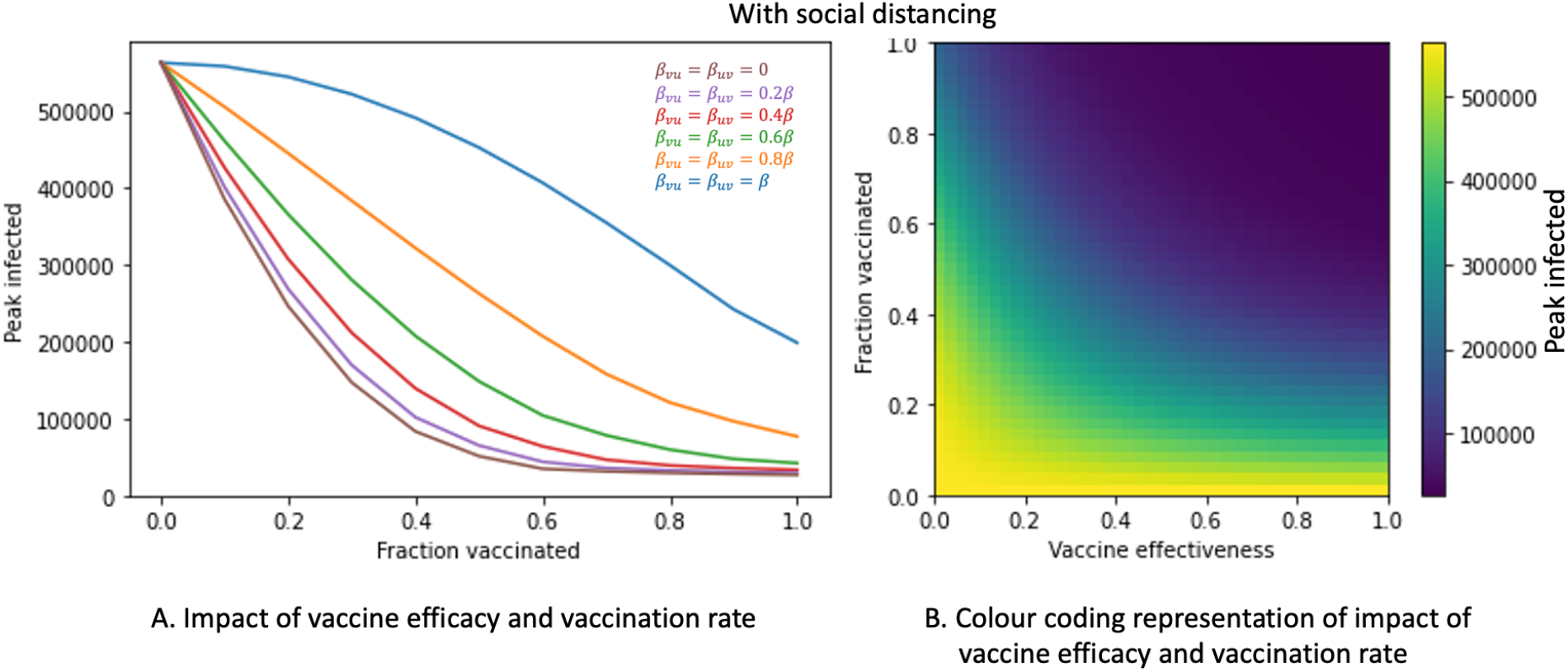
Impact of vaccine efficacy and vaccination fraction on peak COVID infections, with social distancing. A) Impact of vaccine efficacy and vaccination rate. B) Color coding representation of impact of vaccine efficacy and vaccination rate.

In Figure 8(A), peak infected decreases with vaccination fraction for all vaccinated transmission probability values. The decreases in peak infected for lower vaccinated transmission probabilities are sharper than those for higher vaccinated transmission probabilities. As vaccinated transmission probability reflects vaccine efficacy, these results suggest that increasing both the vaccine efficacy and the vaccination fraction helps to decrease peak infections. This conclusion is shown in the color coding scheme in Figure 8(B). As we move from the lower left corner (low vaccine efficacy and a low vaccination fraction) to the upper right corner, peak COVID infections decrease.

Figure 9 represents results of the same model but with the addition of social distancing. Comparing Figure 9(A) to Figure 8(A), it is clear that social distancing slightly shifts the curves for the various vaccinated transmission probabilities down, i.e., social distancing slightly decreases peak infections.

Figures. 8 and 9 demonstrate that peak infections at *κ* = 0 are, for all *ξ* values (again, including *ξ* = 0), significantly larger than the peak infections at *κ* = 1. The color-bars on the right depict the changing peak infections in varying intensities of color as both *κ* and *ξ* are varied.

Figure 10 is, like Figure 7, the overlaid product of two thresholding simulations, but for peak infected. The first (dark blue) shades all (*κ, ξ*) combinations for which the peak infections meet the thresholding condition (peak infections > 5%) in the ‘distancing’ simulation (estimated at 850,000 infected), and the second (light blue) shades the same combinations meeting the same thresholding condition in the ‘no distancing’ simulation. Like the thresholding simulations for total COVID deaths, this overlaid thresholding could be used to observe and compare target reductions. In this case, it seems that the difference between the two thresholds is minute, occurring only when a vaccine is around 95-97.5% effective and 52.5-55% of the population has been vaccinated.

**Figure 10.**
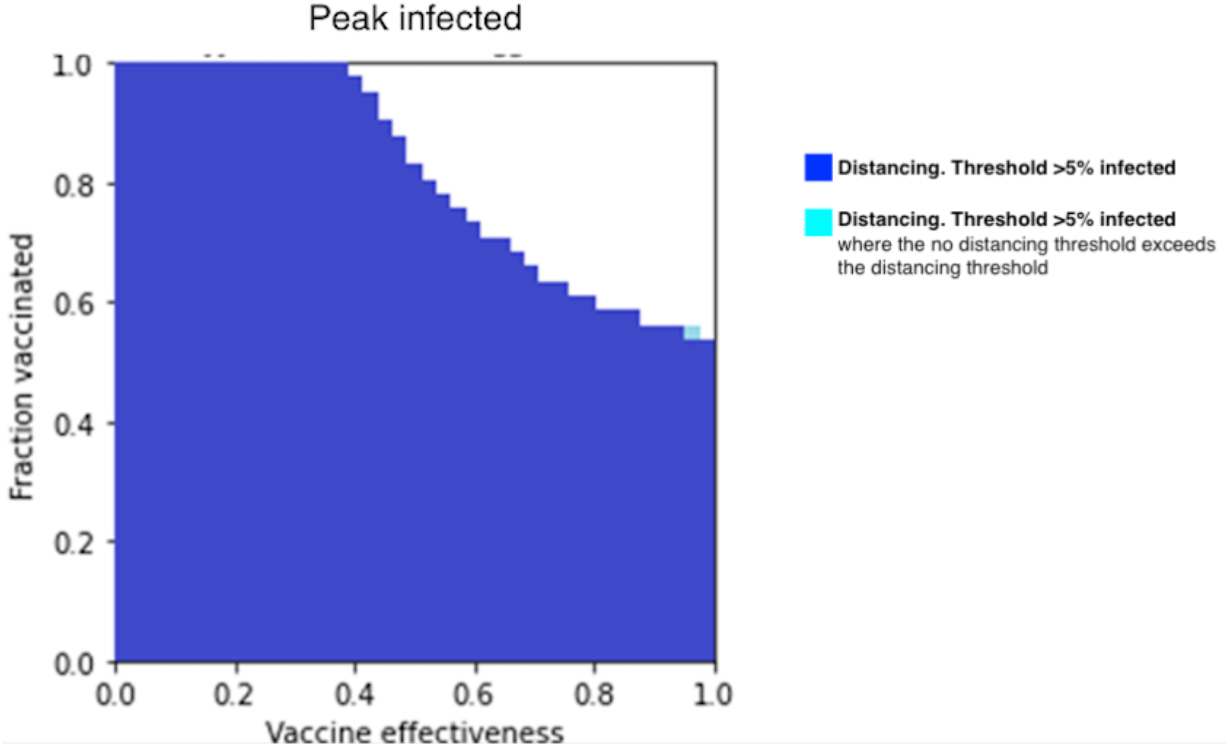
Thresholding of peak COVID infections, with social distancing vs. without social distancing. This figure is similar to Figure. 7, but with the study target being peak COVID infections.

## 4 Discussion

In this study we created a new model to assess the impact of vaccination prevalence, vaccine efficacy, and social distancing on infection rate and mortality from COVID-19. In several countries, including those with developed economies, resources and decision making infrastructure of healthcare systems have proved inadequate during the COVID-19 pandemic, resulting in human suffering and death that could have been mitigated with more informed preparation (25,26). Numerous respiratory pandemics have occurred over the last several decades and have offered opportunities for the scientific and healthcare community to develop effective methods for resource deployment (27,28). Our model presents a tool to inform resource allocation and create anticipatory guidance for public health systems.

The SEIRD+V model we present here advances knowledge in its contribution of new, relevant parameters as inputs: vaccination effectiveness, vaccination fraction, and social distancing stages. These modifications can make the model suitable for different phases of a viral outbreak when the risk-benefit trade-off to calibrate social distancing and other containment policies is shifted. Such insight from a model can help avoid unnecessary social and economic costs. Indeed, during the extended timeline of an epidemic, authorities often need to adapt goals dynamically for public safety in ways that carry a cost to commercial business, education of the young, and general mobility. For example, in the early stages of a pandemic, a common goal for effective containment is to gauge the epidemiological metric R_0_, which depends primarily on infectious period, mode of transmission, and contact rate (29,30). Policy makers may initially seek to limit the contact rate of individuals in a population to a low value, which in the absence of therapeutic options can take the form of policies that promote social distancing. During later stages, authorities may set a goal for vaccine efficacy and vaccination uptake by the population such that no more than a pre-specified fraction of the population is infected. Once vaccines are available, modeling can provide authorities information on vaccine rollout for best results (31–33). This study employs a thresholding method that takes a public health goal as an input and uses it as a threshold to project related conditions necessary to reach such a goal.

Our results show that vaccination drastically reduces peak COVID infections and total deaths from COVID. In a population that has had 50% of its members vaccinated with a 50% effective vaccine, peak infection is reduced from 799 to 161, and maximum total COVID deaths from 30,471 to 8,080 (Figure 4). Our model demonstrates that vaccine fraction in a population decreases mortality, as does vaccine efficacy in every level of vaccination fraction (Figures 5 and 6). These findings corroborate large placebo-controlled trials that show reduced risk of severe disease with vaccination and have additional relevance in the potential to apply such computational analysis to planning and resource allocation for future pandemics (18,19). To explore the impact of social distancing on population health, we analyzed its role in reducing mortality when vaccination fraction was varied (Figures 5 and 6) and found that mortality reduction by social distancing is notably less in the presence of high vaccine efficacy and fraction. Our thresholding results show that more effective vaccines and higher vaccination fractions are required to limit total COVID deaths to the same 5% threshold when there is no social distancing (Figure 7).

Our final set of results deal with the impact of vaccine efficacy, vaccination fractions, and social distancing on peak infections. Total mortality and peak infection rates are reduced by vaccination and social distancing, declining with increased vaccination fractions and with high vaccine efficacy helping to reduce peak infections more sharply (Figure 8). This holds true both when there is social distancing and when there is no social distancing. Social distancing slightly reduces peak infections when the population is vaccinated. Our thresholding results show that vaccine efficacy and vaccination fractions are required at about the same level to limit peak infections to a 5% threshold for both social distancing and no distancing conditions (Figure 10).

Our study is subject to limitations. *First*, a small dataset from a specific locality was used for the simulation, and these data were generated in 2020. SARS-CoV-2 exhibited diverse patterns of spread across the world and even in different counties in the US, with prior studies having developed models, for example, that allow transmission dynamics without peak infection (34). Parameters for transmission and for vaccine efficacy will need to be adjusted for viral subvariants and for evolving vaccination fraction and efficacy in future work. Of note, the model allows for flexible calibration of virus characteristics, which permits refinement of our model with more realistic or more accurate estimates of transition parameters between different compartments. *Second*, the dataset to which this model was applied assessed patients as positive based on a polymerase chain reaction (PCR) saliva-based swab of the mouth that was widely used at the time in Italy. Nasopharyngeal or nasal PCR swabs have become standard in the interval, and the accuracy of the different modalities continues to be studied (35,36). While the dataset may therefore introduce a degree of systematic error, our model can still be useful for assessing relationships between public health goals such as population vaccination rate and effectiveness. Given the internal validity of its results, the model likely also has utility for application to other data cohorts. *Third*, we assumed individuals in each compartment are identical. Numerous studies have shown that individuals differ in outcomes by their age, sex, comorbid burden, and other risk factors (37–39). In the future, this model can be expanded to incorporate time-dependent vaccination, as the current form of this model assumes vaccination as a fixed initial condition. Future versions of the model can also include variables such as percentage vaccination among age groups. *Finally*, we apply assumptions regarding social distancing to allow tractability for modeling. In particular we assume that each individual in the model engages in social distancing to the same extent, and social distancing in our model causes model parameters to change in a uniform way without regard to underlying factors that could change the effectiveness of social distancing. There are various forms of isolation and quarantine that have been developed in professional, educational, and social environments in different countries, and these continue to evolve. Such higher order modeling parameters are beyond the scope of the current work. We sought to offer one form of social distancing in this compartment model to compare its importance to vaccination, which has increasingly become widespread, and found that the impact of distancing likely is attenuated with increased vaccination fraction and efficacy.

In summary, we examined the impact and interactions of social distancing and vaccinations on peak infections and total deaths from COVID-19 using a compartment model and found that vaccination flattens peak infection and reduces mortality, even at low levels of vaccine efficacy and prevalence. Social distancing also decreases peak infection and total mortality when both vaccine efficacy and vaccination prevalence are low, but its benefit is notably attenuated when vaccine efficacy and prevalence are high. Our model’s unique contribution of thresholding to assess vaccine efficacy and vaccination prevalence for preset goals of total mortality and peak infections showed that social distancing noticeably decreases required vaccine efficacy and vaccination prevalence. Tradeoffs between isolation and quarantine on the one hand and vaccine uptake on the other hand will continue to present challenges to decision makers and healthcare professionals. Our model has relevance in its ability to inform social policy decisions and public policy goals in a pandemic that is evolving and continues to pressure us to change our protocols for establishing safe workplace arrangements and sustainable educational environments. It allows for modification of parameters to reflect greater diversity of geography, cultural mores, and viral variants, which is an aim of future work.

## Data Availability

All data produced in the present study are available upon reasonable request to the authors

## 6 Author Contributions

AJ originated the conception and design of the study, created methodology and models, conducted data analysis and interpretation, and drafted the manuscript. HS contributed to conception and design of the study, data analysis and interpretation, and drafting of the manuscript.

## 7 Acknowledgments

The authors would like to thank Jason Leonard for methodological guidance and input and Dr. K. J. Lee for conceptual mentorship throughout all phases of the study.

## References

1. Coronavirus Resource Center [Internet]. Johns Hopkins Coronavirus Resource Center. 2022 [cited 2022 Jul 2]. Available from: https://coronavirus.jhu.edu/

2. CDC. Quarantine & Isolation [Internet]. Centers for Disease Control and Prevention. 2022 [cited 2022 Jul 11]. Available from: https://www.cdc.gov/coronavirus/2019-ncov/your-health/quarantine-isolation.html

3. Hethcote HW. The Mathematics of Infectious Diseases. SIAM Rev. 2000 Jan;42(4):599–653.

4. Brauer F. Mathematical epidemiology: Past, present, and future. Infect Dis Model. 2017 Feb 4;2(2):113–27.

5. Lofgren ET, Halloran ME, Rivers CM, Drake JM, Porco TC, Lewis B, et al. Mathematical models: A key tool for outbreak response. Proc Natl Acad Sci USA. 2014 Dec 23;111(51):18095–6.

6. Chowell G, Sattenspiel L, Bansal S, Viboud C. Mathematical models to characterize early epidemic growth: A review. Phys Life Rev. 2016 Sep;18:66–97.

7. Biggerstaff M, Slayton RB, Johansson MA, Butler JC. Improving Pandemic Response: Employing Mathematical Modeling to Confront Coronavirus Disease 2019. Clinical Infectious Diseases. 2022 Mar 9;74(5):913–7.

8. Carlson CJ, Albery GF, Merow C, Trisos CH, Zipfel CM, Eskew EA, et al. Climate change increases cross-species viral transmission risk. Nature. 2022 Apr 28;1–1.

9. Gibb R, Redding DW, Chin KQ, Donnelly CA, Blackburn TM, Newbold T, et al. Zoonotic host diversity increases in human-dominated ecosystems. Nature. 2020 Aug;584(7821):398–402.

10. Marani M, Katul GG, Pan WK, Parolari AJ. Intensity and frequency of extreme novel epidemics. Proceedings of the National Academy of Sciences. 2021 Aug 31;118(35):e2105482118.

11. Athithan S, Ghosh M. Mathematical modelling of TB with the effects of case detection and treatment. Int J Dynam Control. 2013 Sep 1;1(3):223–30.

12. Zha W, Pang F, Zhou N, Wu B, Liu Y, Du Y bing, et al. Research about the optimal strategies for prevention and control of varicella outbreak in a school in a central city of China: based on an SEIR dynamic model. Epidemiology & Infection [Internet]. 2020 ed [cited 2022 Jul 2];148. Available from: https://www.cambridge.org/core/journals/epidemiology-and-infection/article/research-about-the-optimal-strategies-for-prevention-and-control-of-varicella-outbreak-in-a-school-in-a-central-city-of-china-based-on-an-seir-dynamic-model/FA47CA846564C83234BB95CB5B328278

13. Ram V, Schaposnik LP. A modified age-structured SIR model for COVID-19 type viruses. Sci Rep. 2021 Jul 26;11(1):15194.

14. Cooper I, Mondal A, Antonopoulos CG. A SIR model assumption for the spread of COVID-19 in different communities. Chaos, Solitons & Fractals. 2020 Oct 1;139:110057.

15. Storlie CB, Pollock BD, Rojas RL, Demuth GO, Johnson PW, Wilson PM, et al. Quantifying the Importance of COVID-19 Vaccination to Our Future Outlook. Mayo Clinic Proceedings. 2021 Jul 1;96(7):1890–5.

16. Dashtbali M, Mirzaie M. A compartmental model that predicts the effect of social distancing and vaccination on controlling COVID-19. Sci Rep. 2021 Apr 14;11(1):8191.

17. Carcione JM, Santos JE, Bagaini C, Ba J. A Simulation of a COVID-19 Epidemic Based on a Deterministic SEIR Model. Front Public Health. 2020 May 28;8:230.

18. Polack FP, Thomas SJ, Kitchin N, Absalon J, Gurtman A, Lockhart S, et al. Safety and Efficacy of the BNT162b2 mRNA Covid-19 Vaccine. New England Journal of Medicine. 2020 Dec 31;383(27):2603–15.

19. Thomas SJ, Moreira ED, Kitchin N, Absalon J, Gurtman A, Lockhart S, et al. Safety and Efficacy of the BNT162b2 mRNA Covid-19 Vaccine through 6 Months. N Engl J Med. 2021 Nov 4;385(19):1761–73.

20. Mwalili S, Kimathi M, Ojiambo V, Gathungu D, Mbogo R. SEIR model for COVID-19 dynamics incorporating the environment and social distancing. BMC Res Notes. 2020 Jul 23;13:352.

21. CDC. Coronavirus Disease 2019 (COVID-19) [Internet]. Centers for Disease Control and Prevention. 2020 [cited 2022 Sep 21]. Available from: https://www.cdc.gov/coronavirus/2019-ncov/science/science-briefs/vaccine-induced-immunity.html

22. Kissler SM, Fauver JR, Mack C, Tai CG, Breban MI, Watkins AE, et al. Viral Dynamics of SARS-CoV-2 Variants in Vaccinated and Unvaccinated Persons. New England Journal of Medicine. 2021 Dec 23;385(26):2489–91.

23. Dati COVID-19 Italia [Internet]. Presidenza del Consiglio dei Ministri - Dipartimento della Protezione Civile; 2022 [cited 2022 Jul 2]. Available from: https://github.com/pcm-dpc/COVID-19

24. Berton Ocampo Francisca. Lombardy dataset [Internet]. Available from: https://github.com/pcm-dpc/COVID-19

25. Walker PGT, Whittaker C, Watson OJ, Baguelin M, Winskill P, Hamlet A, et al. The impact of COVID-19 and strategies for mitigation and suppression in low- and middle-income countries. Science. 2020 Jul 24;369(6502):413–22.

26. Flaxman S, Mishra S, Gandy A, Unwin H, Coupland H, Mellan T, et al. Report 13: Estimating the number of infections and the impact of non-pharmaceutical interventions on COVID-19 in 11 European countries [Internet]. Imperial College London; 2020 Mar [cited 2022 Jul 10]. Available from: http://spiral.imperial.ac.uk/handle/10044/1/77731

27. Cauchemez S, Fraser C, Van Kerkhove MD, Donnelly CA, Riley S, Rambaut A, et al. Middle East respiratory syndrome coronavirus: quantification of the extent of the epidemic, surveillance biases, and transmissibility. The Lancet Infectious Diseases. 2014 Jan;14(1):50–6.

28. Donnelly CA, Ghani AC, Leung GM, Hedley AJ, Fraser C, Riley S, et al. Epidemiological determinants of spread of causal agent of severe acute respiratory syndrome in Hong Kong. The Lancet. 2003 May;361(9371):1761–6.

29. Wu JT, Leung K, Leung GM. Nowcasting and forecasting the potential domestic and international spread of the 2019-nCoV outbreak originating in Wuhan, China: a modelling study. The Lancet. 2020 Feb;395(10225):689–97.

30. Ridenhour B, Kowalik JM, Shay DK. Unraveling R0: Considerations for Public Health Applications. American Journal of Public Health. 2014;104(2):10.

31. Moore S, Hill EM, Tildesley MJ, Dyson L, Keeling MJ. Vaccination and non-pharmaceutical interventions for COVID-19: a mathematical modelling study. The Lancet Infectious Diseases. 2021 Jun 1;21(6):793–802.

32. Bubar KM, Reinholt K, Kissler SM, Lipsitch M, Cobey S, Grad YH, et al. Model-informed COVID-19 vaccine prioritization strategies by age and serostatus. Science. 2021 Feb 26;371(6532):916–21.

33. Viana J, van Dorp CH, Nunes A, Gomes MC, van Boven M, Kretzschmar ME, et al. Controlling the pandemic during the SARS-CoV-2 vaccination rollout. Nat Commun. 2021 Jun 16;12(1):3674.

34. Menda K, Laird L, Kochenderfer MJ, Caceres RS. Explaining COVID-19 outbreaks with reactive SEIRD models. Sci Rep. 2021 Dec;11(1):17905.

35. Butler-Laporte G, Lawandi A, Schiller I, Yao M, Dendukuri N, McDonald EG, et al. Comparison of Saliva and Nasopharyngeal Swab Nucleic Acid Amplification Testing for Detection of SARS-CoV-2: A Systematic Review and Meta-analysis. JAMA Intern Med. 2021 Mar 1;181(3):353.

36. Teo AKJ, Choudhury Y, Tan IB, Cher CY, Chew SH, Wan ZY, et al. Saliva is more sensitive than nasopharyngeal or nasal swabs for diagnosis of asymptomatic and mild COVID-19 infection. Sci Rep. 2021 Dec;11(1):3134.

37. Guan W jie, Ni Z yi, Hu Y, Liang W hua, Ou C quan, He J xing, et al. Clinical Characteristics of Coronavirus Disease 2019 in China. N Engl J Med. 2020 Apr 30;382(18):1708–20.

38. Huang C, Wang Y, Li X, Ren L, Zhao J, Hu Y, et al. Clinical features of patients infected with 2019 novel coronavirus in Wuhan, China. The Lancet. 2020 Feb;395(10223):497–506.

39. Mason KE, Maudsley G, McHale P, Pennington A, Day J, Barr B. Age-Adjusted Associations Between Comorbidity and Outcomes of COVID-19: A Review of the Evidence From the Early Stages of the Pandemic. Front Public Health. 2021 Aug 6;9:584182.

